# Labour Care Guide implementation as a decision-making tool for monitoring labour among health care providers in Southwestern Uganda: A protocol for a multisite Effectiveness-Implementation study

**DOI:** 10.1101/2023.06.06.23291028

**Authors:** GR Mugyenyi, EM Mulogo, W Tumuhimbise, EC Atukunda, M Kayondo, J Ngonzi, J Byamugisha, F Yarine

## Abstract

**Background:** The new WHO Labour Care Guide, also regarded as the “next-generation partograph” basing on recent evidence has been recorded as a core component of the 2018 consolidated set of guidelines on intrapartum care for positive child birth experience. The Ugandan Ministry of Health is in the process of adopting the new WHO Labour Care Guide (LCG) with no local context specific data to inform this transition. This study will employ evidence-based research frameworks to identify challenges, and potential opportunities that would inform and refine the implementation strategy and scale-up of this highly promising LCG. We will also seek to utilize best practices to evaluate implementation effectiveness of the new LCG, through employing measurable implementation matrices (implementation, service, patient outcomes).

**Methods:** This study will be a multisite effectiveness-implementation study across all basic and comprehensive emergency obstetric and newborn care facilities in Mbarara district and Mbarara City in Southwestern Uganda. We will employ both quantitative and qualitative methods to evaluate the use of the newly recommended WHO Labor Care Guide in monitoring labor among all health care providers actively engaged in deliverying women across all public maternity health facilities in Mbarara district and Mbarara City. No participant has been recruited at hie point in time

**Results and Discussion:** This study will offer an opportunity to ascertain whether the new WHO Labour Care Guide tool is an effective decision-making tool to monitor labor among healthcare providers conducting routine deliveries in publicly funded facilities in Southwestern Uganda. We will also identify practical, context-specific and actionable strategies for achieving optimal implementation effectiveness in a rural low resource setting.

## Background

The global maternal and neonatal death rates are unacceptably high [1-3] and these deaths are due to complications of pregnancy and childbirth, mostly from preventable or treatable causes. Most of these deaths (94%) are within low- and middle-income countries and could be prevented through timely interventions. In Uganda, the maternal and neonatal mortality remains high at 336 deaths per 100,000 live births and 27 deaths per 1,000 live births respectively [4]. Most maternal deaths in Uganda are linked to prolonged labour; 90% of perinatal mortality following birth asphyxia is directly attributed to obstructed labour [5, 6]. Adequate labour monitoring, with early identification of complications and their management are vital processes towards improved quality of care, and averting the unfavorable delivery outcomes including fetal, new born and maternal deaths [7].

Friedman’s partograph of 1954, also referred to as *the partogram*, is a labour progress/ monitoring chart, graphically depicting the dilatation of the cervix or presenting the dilatation of the cervix against time in labour (Figure 1). Alert and action lines were later added on this Friedman concept by Philpott and Castle in 1972 and aimed at identifying deviations from normal and guiding users towards early intervention. This partogram has been the gold standard for monitoring labour globally [8], and safe motherhood initiatives of the 1990s rolled it out as a universal tool for monitoring labour in an effort to prevent prolonged and obstructed labour. However, despite decades of health care provider (HCP) training, support and investment, rates of partograph utilization, acceptability and appropriate use in making critical decisions during labour remain sub optimal in resource-limited settings, with correspondingly high incidences of obstructed labour, its associated complications, and ultimately sustained high and unacceptable still birth, maternal, and neonatal mortalities [1, 9, 10] [11] [9, 12]. The partogram also offers subjective variations, and assumes that all women progress at same rate, which may affect intervention rate [13]. A study of 527 parturients in Southwestern Uganda observed that 77.6% of clinical records actually contained a partograph, and an abysmal 4.2% had been completed to standard [12]. In fact, according to Lugobe and colleagues, the partograph was most commonly used to record birth outcomes and not the actual monitoring of labour which it is intended.

**Figure 1:**
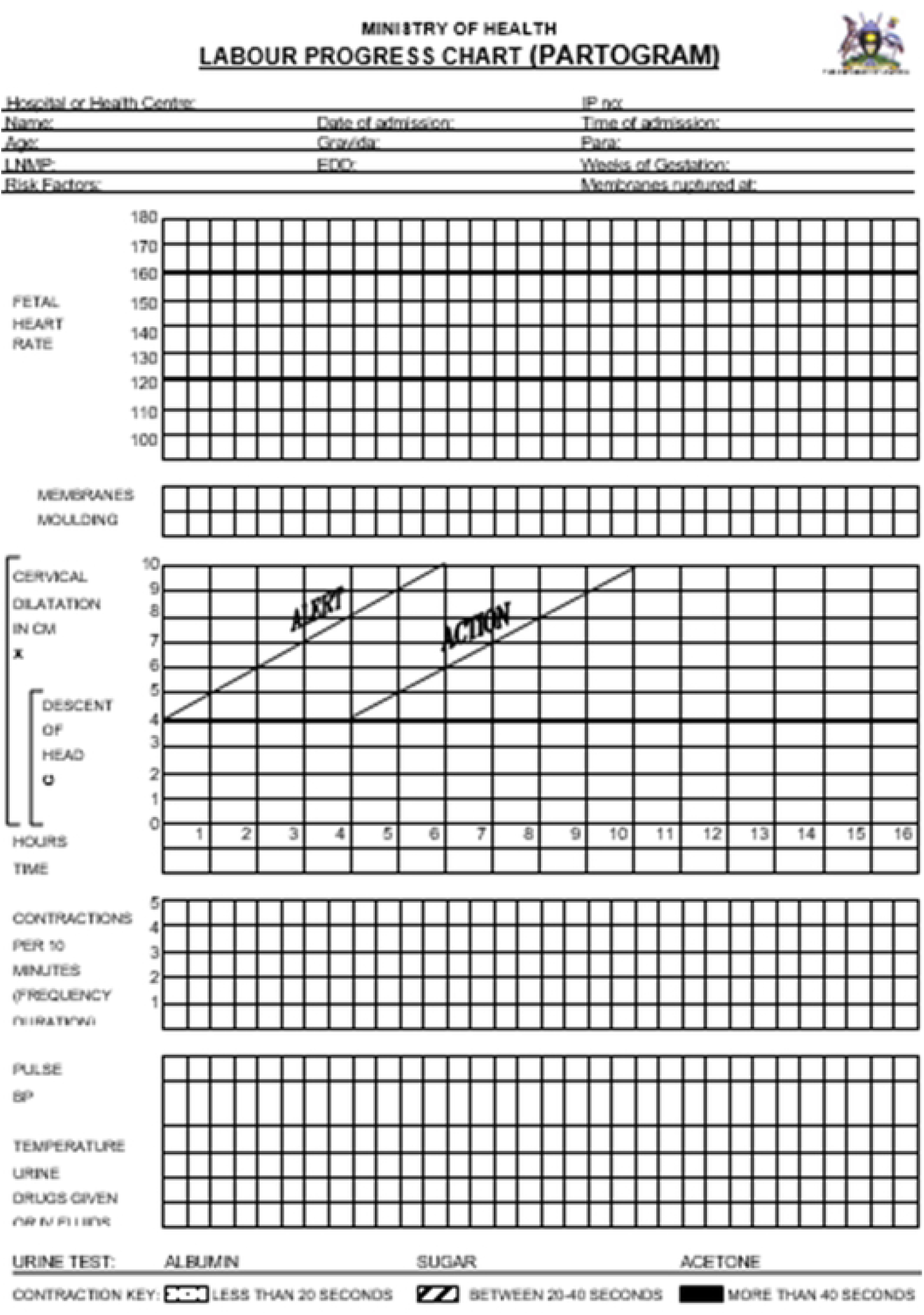
The Labour progress chart (Partogram)

Use of effective evidence-based interventions during labour, childbirth, while avoiding ineffective or potentially harmful ones could facilitate all women to achieve desired emotional, psychological and physical outcomes through regular assessments to identify any deviation from normality (WHO, 2020). With the persistent maternal/perinatal mortalities, the Uganda’s Ministry of Health (MOH) has launched the new Essential Maternal and Newborn Clinical Care (EMNC) guidelines for Uganda in which the reproductive health experts recommended replacing the partograph with the new WHO Labor Care Guide (LCG) [5]. This recommendation was made with anticipation that the new tool was easy to use, promptly identifies deviations from normal through regular assessments, encourages self-efficacy, stimulates HCP interaction and shared decision making, recognizes participation of labour companions to promote women centered care, but importantly the LCG has been developed for health care providers to identify deviations from normal through regular monitoring and assessment of women and their unborn babies. This LCG has been encouraged in place of the partograph to monitor the well-being of women and babies during labor to identify any deviation from normality (WHO, 2020). The LCG tool encourages health care team interaction, and aims at initiating or stimulating interaction among health workers, women and companion/family members, while emphasizing safety, and providing evidence based supportive care which has been found to be paramount among expectant women in our setting (Atukunda et al., 2020), while avoiding unnecessary practices, and solve the sustained challenges that were faced by the HCPs while using the partograph over the decades (user-centered) (WHO, 2020). In fact, according to Vogel and colleagues, this new WHO labour care guide monitoring tool is regarded as the “next-generation” partograph incorporating recent effective intrapartum care guidelines [14]. When used properly the LCG is thought to effectively detect prolonged labor in time for HCPs to perform required interventions in time before progressing to obstructed labor and its sequalae; such as ruptured uterus, post-partum hemorrhage, sepsis, maternal and neonatal deaths. For example, while the partograph lacked clearly defined identifiers of prolonged/obstructed labour, the new LCG identifies grade three moulding and caput, all represented by +++, and further defines prolonged labour using cervical dilatation specific time lags indicated in its “alert” column of section 5 (Figure 2) as per the new WHO consolidated guidelines on intrapartum care for positive childbirth [15]. In fact, the new LCG has alerts for all observations that prompt the labour monitoring team to take action on each abnormal observation as soon as it happens, rather than waiting for the plot to reach action line as it is in the partograph.

**Figure 2:**
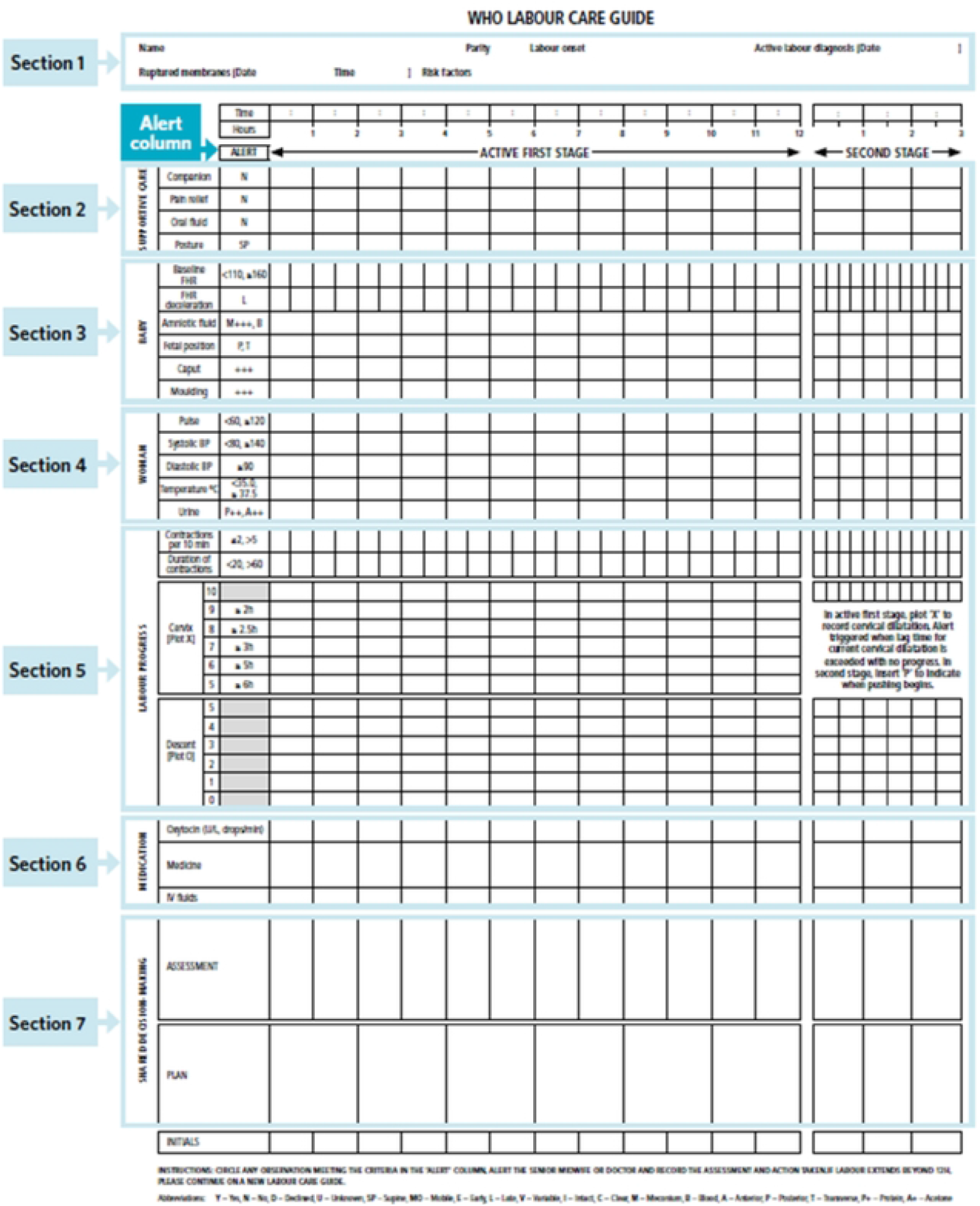
The WHO Labour care Guide.

Uganda’s MOH is in the process of rolling out the implementation of the LCG to all health facilities. However, there is no local context specific data to inform this transition. Many new interventions have failed because of inattention to implementation needs early during their development. This study will employ evidence-based research frameworks to evaluate the effectiveness and implementation process of LCG, through employing measurable implementation matrices (implementation, service, patient outcomes). This will help to identify potential opportunities, challenges, inform and refine the implementation strategy and scale-up of this highly promising LCG. We will utilize best practices to develop a context specific tool that is aimed at improving end-user (HCPs) acceptability, satisfaction, motivation, appropriateness, feasibility, fidelity, patient-centeredness and effectiveness of this new tool in monitoring labour progress in a rural Southwestern Ugandan community setting where the impact of such an intervention is likely to be the greatest.

## Methods

### Study design

We shall conduct a mixed method multisite effectiveness-implementation study across public health facilities in Mbarara district and Mbarara City -Southwestern Uganda to refine the newly recommended WHO Labor Care Guide in monitoring labor during routine care and evaluate its effectiveness and implementation using Proctor’s implementation outcomes framework as outlined in Table 1. We hypothesize that the modified LCG will improve labour monitoring, reduce prolonged labour, obstructed labour, its complication, ultimately reducing the maternal and neonatal deaths. The proposed interviews will help to refine implementation strategies for scale-up using the Consolidated Framework for Implementation Research (CFIR) as outlined in Table 2. Our outcomes will also serve as indicators of implementation success or necessary pre-conditions for attaining desired service outcomes for HCPs carrying out deliveries in rural, resource-limited settings.

**Table 1:** Application of the Proctor framework to evaluate acceptability, appropriateness, user satisfaction, feasibility and fidelity.

| Domain | Definition | Specific intervention measures |  |
| --- | --- | --- | --- |
| Acceptability | Agreeability, palatability, complexity, comfort, experience | -Reported ease of use/performance expectancy, tool engagement<br>-Effort expectancy, facilitating conditions, social influence<br>-Self-efficacy, behavioral intention to use LCG/intervention<br>-I will measure acceptability using Weiner's tool, [33, 34] | -Exit interviews<br>-Questionnaire |
| Adoption |  | -Initiation and use of the LCG over time<br>-% completed LCG over time | -Document review |
| Satisfaction | Satisfaction with content, delivery & credibility | -Satisfaction (Health care provider Satisfaction Questionnaire; CSQ-8) [35] | -Exit interviews<br>-Questionnaire |
| Appropriateness | Perceived innovation fit, relevance (for setting), compatibility | -I will measure appropriateness using Weiner's tool, [33, 34]<br>-Relevance for setting, compatibility | -Questionnaire<br>-Exit interviews |
| Feasibility | Extent to which the new *labor care guide* can be successfully used within a given setting | -Proportion of HCPs willing to take up the LCG, recruitment rates, reason for not participating, user demographics, retention, participation/complete utilization rates, Proportion of HCPs able to fully fill the LCG, prompts responded to on the LCG, time taken to respond, proportion of unnecessary interventions, proportions of actions taken on time<br>-I will also use Weiner's tool, [33, 34] to measure feasibility | -Exit interviews<br>-Document review<br>-Questionnaire |
| Fidelity | -Adherence to protocol (extent to which intervention was delivered as intended)<br>- Exposure to intervention (participant involvement & responsiveness)<br>-Quality of delivery | -Percentage of fully filled LCGs<br>-Proportion of HCPs with capacity/integrity to deliver LCG appropriately (adherence to protocol, timeliness, cues to action, motivation, HCP Vs patient number ratio)<br>-Awareness, skills to deliver, involvement, responsiveness<br>-Quality of program delivery- LCG training, comprehension, efficiency in decision making<br>-Number of prompts responded to, engagement with other staff, average length of 1<sup>st</sup> and second stage of labor<br>-Clarity on the LCG<br>-Number and type of technical issues/ problems encountered (no staff to assist/ respond, etc)<br>-I will adopt the support fidelity scale[36] to additionally assess fidelity | -Exit interviews<br>-Document review<br>-Questionnaire<br>-Clinic data |
| Effectiveness | See objective 3 | -See objective 3 |  |
| Penetration | Level of institutionalization, service access | -Number and type of HC/HCPs consistently utilizing the LCG over time,<br>-HCP motivation to sustained LCG use | -Facility Audits<br>-Exit interviews |
| Sustainability | Integration, sustained use, routinization, institutionalization, continuation, durability | -Consistent & complete use over time (user retention), engagement of staff (shared decision) on the LCG over time, HCP perceived usefulness of the LCG over time | -Document reviews<br>-Exit interviews |
| Efficiency | Ability to achieve sustained LCG use with minimal effort | -Time spent on using the new LCG, timely detection of prolonged labour versus the previous partogram | -Exit interviews<br>-Document review |
| Patient/user-centredness | User driven design/content<br>Ability to put users needs at the center | -Pain relief, inclusion of labor companion, shared decision, customization, contextualization, & tailoring of the LCG (objective 1) | -Exit interviews |
| Timeliness | Impact over time | -Perceived impact on prolonged labour and obstructed labour over time | -Exit interviews |
| Function | Functionality/suitability/practicality | -Perceived quality, impact on labour monitoring & obstetric care | -Exit interviews<br>-Questionnaire |

**Table 2:**
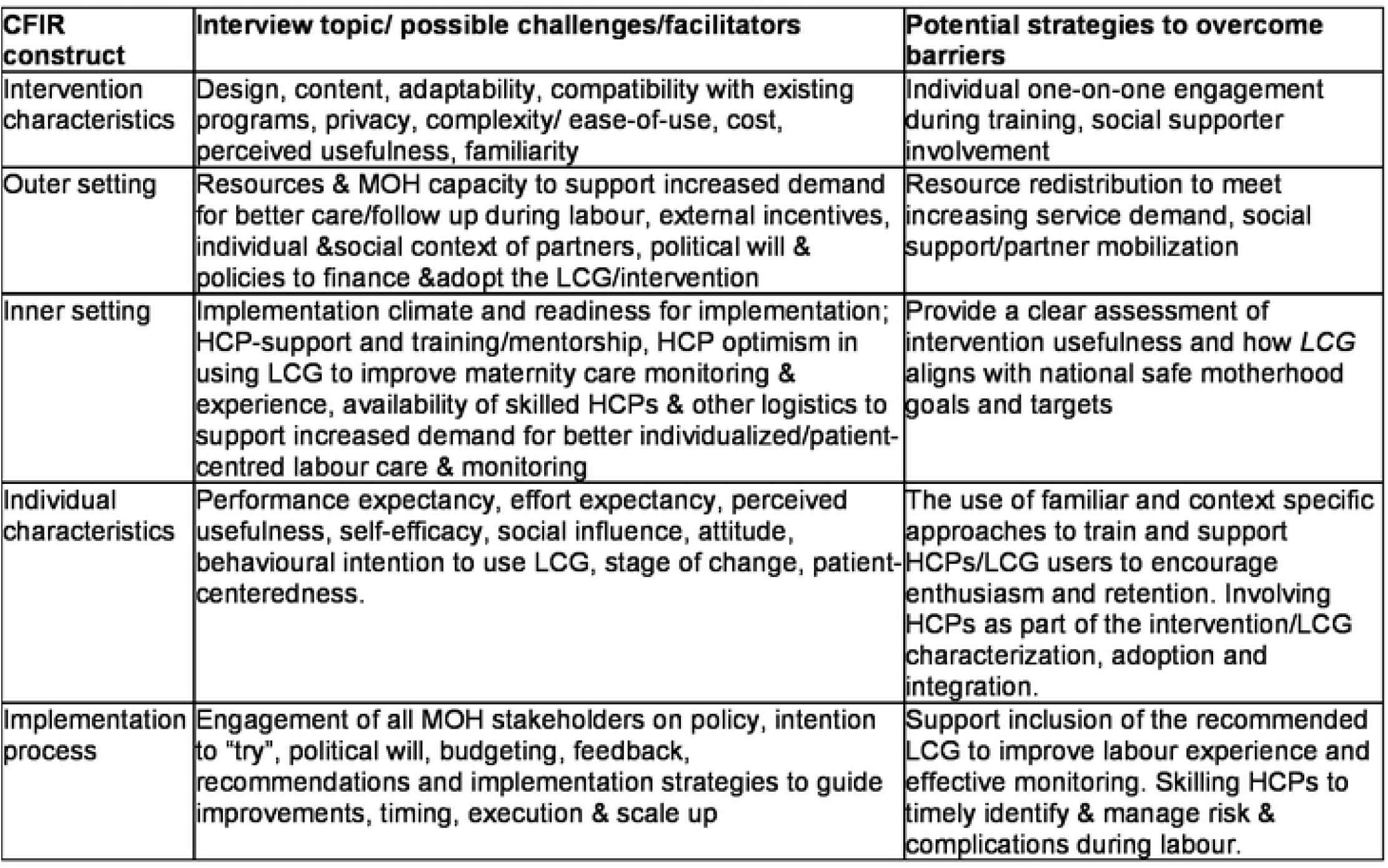
CFIR constructs that will guide data collection on intervention challenges, facilitators, and potential strategies by HCPs and payers/managers.

We will utilize an ambispective cohort; a combination of a historical cohort of mothers monitored using a partograph and prospective cohort of women monitored using the new tool to evaluate implementation success (effectiveness). As a concerted effort to meaningfully implement a new intervention meant to reduce preventable maternal and perinatal morbidity and mortality, these results will generate grounded, robust scientific data to inform stakeholders and policy makers working towards effectively integrating and scaling up of this new LCG into routine maternity care in similar settings across the country and beyond. This study will also be able to show the effect of this intervention, and optimize its implementation in routine maternity care practice to improve maternal-fetal outcomes in similar settings. No participant has been recruited at hie point in time

### Study setting

The study will be carried out in all public health facilities offering basic and comprehensive emergency obstetric and newborn care in Mbarara district and Mbarara City. These include all 11 public Health center threes (HCIII), two Health Center fours (HCIV) (Bwizibwera, Mbarara City Council), and one regional referral Hospital (Mbarara Regional Referral Hospital (MRRH). Mbarara District is located approximately 270 kilometers Southwest of the capital, Kampala, with a population of abount 250,000 people distributed through two recent administrative units of Mbarara City and Mbarara district. Uganda’s public health system is organized into seven tiers with national and regional referral hospitals, general district hospitals and four levels of community health centers. Staffing and available services vary across the four levels: HCIII offer basic emergency obstetric care (carry out ANC and conduct vaginal deliveries), whereas Health Center one (HCI) and Health Center two (HCII) serve as low resource primary health care units. HCIVs and hospitals conduct normal and caesarean deliveries (offer comprehensive emergency obstetric care), and have ambulances and blood transfusion services [16]. Private providers operate in parallel to the public health system to provide maternal health care. Basic and emergency obstetric care services are provided through four hospitals; Mbarara Regional Referral Hospital and four privately owned; Divine Mercy Hospital, Ruharo mission Hospital, Mbarara Community Hospital, Mayanja Memorial Hospital, and with two HC IVs of Bwizibwera and Mbarara City. Mbarara is served by 11 HC IIIs, and over 40 privately-owned health facilities that provide maternity services. The district is served with a total of 253 HCPs who provide obstetric health care with a large concentration at Mbarara Regional Referal Hospital and the health center IVs (2 medical officers and 10 midwives on average) [17].

The local economy of the districts is largely based on subsistence agriculture, with both food and water insecurity being common [18]; ANC attendance of ≥4 visits is still at 58%, and maternity services, including delivery, are largely provided free of charge. The study will be conducted at the labour suite and post-natal wards of all the 11 HCIIIs, (6 from Mbarara district including Bubaare, Bukiro, Kagongi, Kashare, Rubaya and Rubindi distributed in the 6 subcounties of Mbarara District and 5 HCIIIs of Biharwe, Kakoba, Nyakayojo, Nyamitanga and Kyarwabuganda distributed in the 6 didvision of Mbarara City plus 2 HCIVs (of Bwizibwera and Mbarara City) and Mbarara Regional Referral Hospital. All mothers in labour are ideally monitored using a partogram and the fetal heart rate is measured manually per clinician judgment using the Pinard. After a normal (un complicated) vaginal delivery, the mothers from these facilities with their babies are admitted to the post-natal wards for 24 hours, with daily ward rounds conducted by skilled birth attendants. Those who deliver by caesarean section remain admitted for 3 to 5 days though mothers and babies with complications are admitted for more days.

### Study population

All adult HCPs actively involved in maternity care and conducting deliveries, health facility managers in Mbarara district and officials from the reproductive health division of the Ugandan Ministry of Health will be included in this study. The principal investigator will provide a list of these eligible HCPs to the study research assistants, who will then contact and seek written informed consent before enrolment into the study. HCPs will be taken through the study procedures. Eligible local and national MOH facility managers/stakeholders who participate in budgeting, procurements or funding for facilities will also be identified and enrolled into the study.

### Eligibility Criteria

Individuals with self-reported willingness to use the new LCG in monitoring of labour, able and willing to provide informed consent will be invited to participate in this study. Individuals unwilling to use the LCG and unable to provide informed consent will not be eligible to participate in this study.

### Study procedure

We will carry out baseline needs assessment among HCPs who actively engage in deliverying women in Mbarara district and Mbarara City for a period of atleast 1 year before the introduction of the LCG. Guided by the Consolidated Framework for Implementation Research CFIR [19], we will first identify unique needs, challenges, facilitators and patterns of potential and sustained uptake of the new intervention (LCG) at different levels of the ‘4 tier’ health system to monitor labour in rural Southwestern Uganda (Aim 1). This will involve conducting in-depth qualitative interviews with up to 30 purposively selected HCPs, and 15 Ugandan MoH officials at the onset of the study. We will then characterize, and refine the new WHO LCG based on the findings from Aim 1, and then develop a suitable implementation strategy to effectively integrate the new LCG into routine maternity care in Mbarara district and city. We will iteratively test the LCG prototype amongst three sets of 10 HCP users (interviewed from objective 1). The aim is to refine and customize the tool for easy uptake and sustained utilization within a known intervention development framework [20-22]. Ten women in three successive iterations will be involved to test subsequent prototypes. Upon completion of each of the one-week period, we will interview HCPs using structured questionnaires to obtain feedback on the ease of use, complexity, content, tool’s ability to engage, motivate, prepare, request or get support/attention as needed, cues to action/alerts/prompts, social support, guidance on what to do at each stage for optimal and timely response. After each iterative round, we will hold group discussions at the local and MOH level to explore user experiences, define and refine relevant components of the LCG. The final LCG prototype will be ready for evaluation in routine care on a bigger scale (Mbarara district and Mbarara city health facilities).We will also customize the existing training manuals developed by WHO to suite within the local Ugandan context using this feedback. This is aimed at improving the skill, ease-of-use, and appropriate utilization of the developed LCG to maximize impact. Training on LCG use will generally aim at behavioral change communication; reference information and foot notes will be integrated within the labor care guide prints to facilitate exposure, awareness, accurate delivery, usability, comprehension and decision making [20].

Once the refined prototype is completed, we will evaluate the LCG use by HCPs conducting deliveries from all basic and comprehensive emergency obstetric and new born care facilities of Mbarara district and Mbarara City. We will utilize the Proctor implementation outcome framework (Table 2) to evaluate implementation outcomes of using the new labour care guide in routine maternity care that include; acceptability, appropriateness, feasibility, fidelity, and effectiveness among HCPs actively involved in deliveries across Mbarara District and Mbarara City. A data abstraction tool for women who were monitored using the partograph (historical cohort) and prospective data for those monitored using the new LCG will be designed and used in the document review to collect data from patient facility records. We will design a database and enter all partograph and LCG abstracted data. We will document and assess delivery outcomes (effectiveness) such as duration of active phase of labor, duration of second stage of labor, presence of labor companion, initiation of breastfeeding and other relevant maternal-fetal outcome events as described in the data collection section. A chart review of the files and or postnatal discharge forms will also be done to document tool completion, as well as actual labour and delivery outcomes

Finally, we will assess the diagnostic predictability of the new LCG compared to the partogram in effectively detecting prolonged labour and reduce rates of obstructed labour among women delivering in Mbarara district and Mbarara City. We will use the effectiveness data to assess the specificity, sensitivity, negative and positive predictive values of the new LCG versus partogram as the presumed standard of care. Because the key identifiers of obstructed labour were not clearly defined and indicated on the previous partogram, namely grade 3 moulding and caput, we will use prolonged labour as a quantifiable comparative measure. Prolonged labour will be defined using a partogram as labour progress that crossed the action line figure 1), and cervical dilatation specific time lags indicated in the “alert” column when using the LCG. Whereas prolonged labour was defined at the end as labour crossing the action line on the partograph with no ongoing prompts in between, the new LCG recommends on going practical observable “time lag” at each specific centimeter of cervical dilation as specifiied in the “alert” column of section 5 which empowers HCP to make decisions before obstructed labour develops, indicated by grade 3 moulding/caput in section 3 of the new LCG.

We will summarize the clinical and demographic data obtained from document review of the patients in the ambispective cohort. We will then assess the diagnostic validity of the new LCG versus the partogram as the presumed standard of care. To describe the performance of the LCG compared to the partogram, we will consider prolonged labor as a dichotomus outcome to asses the specificity and sensitivity of the LCG, and fit a receiver operating curve at different time points from 4 centimetre cervical dilatation (for partograph) and 5 centimeter cervical (for the new LCG) till vaginal delivery or decision to perform caesarean section.

### Outcomes

The primary effectiveness outcome will be the proportion of women with prolonged labour. We will define prolonged labour as 1) labour crossing the action line on the partograph, 2) labour lasting more than a specified centimeter cervical dilation “time lag” in the alert column of section 5 of the LCG.

Secondary outcomes will include; proportion of obstetric interventions such as caesarean sections, labour augmentation, blood transfusion; quality-of-care; having a fresh still birth; duration of 1^st^ and 2^nd^ stages of labor; 5-minute apgar score, need for rescuscitation/blood transfusion, mode of delivery; initiation of breastfeeding; obstetric complications diagnosed and or managed during labor, childbirth or immediate postpartum; ruptured uterus; postpartum hemorrhage; maternal/newborn sepsis; maternal, fetal, and newborn deaths.

### Power and sample size calculations

This study will enrol all HCPs actively engaged in delivering women across public facilities offering basic and comprehensive obstetric care across Mbarara district. Current statistics indicate a 2% of obstructed labor, and 21% of obstructed labor complications [23]. Consequently, Uganda suffers from one of the highest maternal mortality ratios (336 for every 100,000 women), and child perinatal mortality rates (41 deaths per 1000 births) in the world. The study team will review all the documents one year before and one year after implementation of the LCG to assess effectiveness and implementation success. As we move the LCG to a community setting with more users, subject to local public health facility challenges, uptake, utilization, and training needs may vary [24]. A 10% reduction would therefore be meaningful for policy makers to signify public health importance, and in demonstrating an improved maternal-newborn outcomes at the local community level. Since we are enrolling and reviewing all documents/files, we will have sufficient power to detect secondary maternal-newborn outcomes that would be more informative for maternal health policy makers and implementers. We will therefore have sufficient power to detect a programmatically meaningful 10% superiority reduction rate of prolonged labour/obstructed labour from over 20,000 deliveries recorded across Mbarara district and Mbarara city annually after implementation of the LCG.

### Data collection and Management

The demographic and clinical data will be collected from maternity records of women that have delivered within one year before and a year after implementation of the LCG including; patient demographics (e.g age, gravidity, parity, gestational age), prenatal, antepartum high-risk morbidities, non communicable diseases (NCDs), and HCP demographics; age, education, experience, self-efficancy. All data will be entered into REDCap and checked for completeness and quality by the principal investigator and any problems that arise will be resolved immediately.

Charts of women whose labour was monitored using a partograph one year before introduction of the new WHO LCG will constitute a partogragh-historical cohort (control arm) while those monitored using the new LCG will form the prospective LCG (intervention arm) of the study. We will first summarize health-related and socio-demographic data between arms. For our primary effectiveness outcomes, we will fit a multivariable logistic regression model, with study arm as the predictor of interest, and age, high-risk pregnancy and health facility at enrollment as *a priori* additional variables in the model, due to their strong association with the selected outcome[25-27]. Although not designed to detect a difference, we will also explore additional secondary outcomes, as listed above. We will also summarize implementation outcomes for the new LCG users (HCPs) using descriptive statistics. Success in the implementation survey data will be identified qualitatively and by the top tertile of relevant scales (e.g., acceptability, feasibility, satisfaction, appropriateness). We will also describe the ranked implementation strategies selected by LCG users and key MOH stakeholders observed during the feedback interviews. Lastly, we will calculate the positive and negative predictive values of prolonged labour, and summarize it in relation to the calculated prevalence, as well as the documented prevalence in Uganda. Data analysis will be conducted using STATA version 17 (Statacorp, College Station, Texas, USA). Findings will be presented as descriptive statistics, scatter plots and graphs; statistical significance will be considered at p ≤ 0.05.

### Qualitative analysis

All transcripts from the interviews will be done and transcribed in English by two research assistants. The aim of this analysis will be to inductively construct categories describing multilevel factors and strategies that might influence LCG implementation and effectiveness. Qualitative analysis will be inductive, and a codebook will be developed through conventional content analysis [28]. To ensure accuracy, transcripts will be coded to calculate an intercoder reliability Kappa statistic using the NVIVO software Version 12 (Melbourne, Australia). We will begin the category construction process with repeated review of transcripts to identify relevant content. Identified content will serve as the basis for developing a coding scheme. Coded data will be iteratively reviewed and sorted to suggest categories under the general headings as per the employed CFIR framework. The categories developed from coded data will consist of descriptive labels, elaborating text to define and specify each category’s meaning, and illustrative quotes taken from the qualitative data. Demographic data will be used to describe the sample.

### Ethical considerations

Ethical clearance was obtained from the Faculty Research Committee (FRC) in the faculty of Medicine and the Research Ethics committee (REC) (Protocol number: MUST-2023-808) at Mbarara University of Science and Technology. Study site administrative permission was obtained from Mbarara Regional Referral Hospital, Mbarara District Health officer and the City Health Officer for Mbarara City. We obtained approval from the National Council for Science and Technology in Uganda and will obtain written informed consent from all study participants before enrolment in the study.

### Publication and dissemination of results

The research outcomes from this study will be published in international peer reviewed open access journals and the Ugandan Ministry of Health, and selected national and international conferences. The study will be registered with clinicaltrials.gov.

## Discussion

This study offers an opportunity to ascertain whether the new LCG tool is an effective decision-making tool to monitor labor among obstetric care providers in publicly funded facilities in rural settings such as Uganda. The WHO LCG has been referred to as a “next-generation” partograph for HCPs in adequately monitoring the well-being of women and babies during labour and childbirth, timely identifying any deviation from normal and facilitating HCP interaction, stimulate shared decision-making for HCPs, laboring women and their companions/family thus facilitating timely management, quality of women-centered care and birthing experience. The tool aims toThe reference thresholds for abnormal labour observations (for each monitored parameter) provided by the new LCG are meant to trigger specific actions, and thus targets to minimizeover-diagnosis and under-diagnosis of abnormal labour events and the unnecessary use of interventions such as caesarean sections and augmentation.

### Limitations

This study will be conducted at a time when the Ugandan MOH will be recalling the use of a partogram and with some isolated facilities already using the original WHO LCG. We will therefore be unable to conduct a well-controlled prospective cohort using the obsolete partograph, hence our choice to utilize a historical or retrospective cohort of partograph data to conduct this study. While we may find more missing data in the partograms for the retrospective partograph arm, we have powered the study to deal with the missing data and entering data from all available partographs at the facilities for a whole year proceeding the new LCG introduction. However, in sensitivity analyses, we will repeat the analysis after excluding women with missing outcome data.

### Implication to policy makers and implementers

No rigorous adoption and evaluation of the effectiveness and implementation of the new WHO LCG intervention has been conducted in Uganda or similar setting to inform stakeholders on successful roll out of the intervention on a large scale, and long term. The Ugandan Ministry of Health is in the process of adopting the new WHO LCG with no local context specific data to inform this adoption and transition for use in routine care. This study therefore seeks to utilize best practices to support intervention adoption, uptake, implementation, integration and scale up. This study will offer an opportunity to ascertain whether the new WHO Labour Care Guide tool is an effective decision-making tool to monitor labor among healthcare providers conducting routine deliveries in publicly funded facilities in Southwestern Uganda. We will also identify practical, context-specific and actionable strategies for achieving optimal implementation effectiveness in a rural low resource setting. This study has potential to prevent a substantial number of annual 823,000 stillbirths, 1,145,000 neonatal deaths and 166,000 maternal deaths in the 75 highest burden countries [29-32].

## Data Availability

All relevant data from this study will be made available upon study completion.

## Notes

### Competing Interest Statement

The authors have declared no competing interest.

### Funding Statement

The author(s) received no specific funding for this work

### Author Declarations

Ethical clearance was obtained from the Research Ethics committee (REC) (Protocol number: MUST-2023-808) at Mbarara University of Science and Technology. Study site administrative permission was obtained from Mbarara Regional Referral Hospital, Mbarara District Health officer and the City Health Officer for Mbarara City. We obtained approval from the National Council for Science and Technology in Uganda (HS2864ES) and will obtain written informed consent from all study participants before enrolment in the study. No data has been collected so far

